# Quantifying the Effects of Social Distancing on the Spread of COVID-19

**DOI:** 10.1101/2020.09.19.20197996

**Authors:** Talal Daghriri, Ozlem Ozmen

## Abstract

This paper studies the interplay between the social distancing and the spread of COVID-19 disease—a widely spread pandemic that has affected nearly most of the world population. Starting in China, the virus has reached the United States of America with devastating consequences. Other countries severely affected by the pandemic are Brazil, Russia, United Kingdom, Spain, India, Italy, and France. Even though it is not possible to eliminate the spread of the virus from the world or any other country, it might be possible to reduce its effect by decreasing the number of infected people. Implementing such policies needs a good understanding of the system’s dynamics, generally not possible with mathematical linear equations or Monte Carlo methods because human society is a complex adaptive system with complex and continuous feedback loops. As a result, we use agent-based methods to conduct our study. Moreover, recent agent-based modeling studies for the COVID-19 pandemic show significant promise assisting decision-makers in managing the crisis through applying some policies such as social distancing, disease testing, contact tracing, home isolation, providing good emergency and hospitalization strategies, and preventing traveling would lead to reducing the infection rates. Based on imperial college modeling studies that prove increasing levels of interventions could slow down the spread of disease and infection cases as much as possible, and taking into account that social distancing policy is considered to be the most important factor that was recommended to follow. Our proposed model is designed to test if increasing the social distancing policies strictness can slow down the spread of disease significantly or not, and find out what is the required safe level of social distancing. So, the model was run six times, with six different percentages of social distancing with keeping the other parameters levels fixed for all experiments. The results of our study show that social distancing affects the spread of COVID19 significantly, where the spread of disease and infection rates decrease once social distancing procedures are implemented at higher levels. Also, the behavior space tool was used to run ten experiments with different levels of social distancing, which supported the previous results. We concluded that applying and increasing social distancing policy levels led to significantly reduced infection rates, which result in decreasing deaths. Both types of experiments prove that infection rates are reduced dramatically when the level of social distancing intervention is implemented between 80% to 100%.

## I. Introduction

In 2020, the novel coronavirus (COVID-19) has spread across the globe, taking lives and wreaking havoc. The world has not seen a health threat like this since the 1918 influenza pandemic. During that pandemic, at least 50 million people died, and one-third of the world’s population was affected. The full impact of the coronavirus pandemic is yet to be seen as cases continue to rise exponentially. Since there is not yet a vaccine for COVID-19, a lot of uncertainty remains about its impact and the best way to slow the spread of the virus. Countries around the world are implementing various policies such as social distancing, which aims to reduce the amount of contact people have with each other.

The effectiveness of various policies is best shown through data collected from China and Italy, which were the first countries to feel the impacts of the pandemic. Italy was slower to implement social distancing policies than China, and it is evident from this data that they paid the price for their inactivity. Meanwhile, the economic cost can be seen in the data from China and from the US, where there have been unprecedented numbers of people claiming unemployment benefits. Goldman Sachs predicted quarter-on-quarter annualized growth rates of -6%in Q1 and -24% in Q2 for the US [1]. It clear that social distancing policies are taking their toll on the economy and the future looks bleak.

With such high economical costs and long-term impacts of strict social distancing policies, questions about whether such policies are worth it will naturally arise. The data coming out of China (as well as South Korea and Singapore) demonstrates the effectiveness of social distancing at slowing the spread of coronavirus; however, it is possible that less strict interventions would have a less severe impact on the economy while still slowing the spread [2][3][4].

At this point, the main study that has examined the data to weigh up the benefits of social distancing versus the economic cost compared two scenarios [1][5]. The first scenario is 3-4 months of social distancing in the form of isolating people with symptoms at home, quarantining at home people who live in the same house as the suspected case and social distancing for the elderly and people most at risk. The second scenario is without any social distancing. This study showed that in the first scenario (social distancing) there would be 1.76 million fewer fatalities from COVID-19 over a period of 6 months than in the second scenario (no policy).

We developed an agent-based simulation model by using Netlogo simulation software to quantify the effect of social distancing on the spread of COVID-19 disease. Simulation helps in conducting our study since we can replicate the case study with much less cost and predict valid outcomes that help in providing the right recommendations to save the lives and improve the required health care services during the crisis. Our simulation model can be verified and validated with the data from the countries that have implemented social distancing and those countries which cannot implement due to any reason. It would certainly help the policymakers to decide what to do and what not to do. It is intended to be a decision support system that can show the outcome effectively and efficiently. In our study, we seek to quantify the safest level of social distancing that should be applied and followed by the citizens in the United States.

## II. METHODS

We used agent-based modeling to capture the cause-effect relationship of COVID-19 with the well-known intervention of social distancing. We used ODD+ D protocol, which is an extension of the ODD protocol, where ODD+ D means (Overview, Design Concepts, and Details) + decision. Moreover, ODD+ D was extended to introduce human decision behavior to the previous code so it would help to study and analyze the human behaviors regarding the decision-making process [6] [7].

### A. Overview

1. Purpose: This paper describes the development and implementation of a simulation model in NetLogo software to understand the effect of social distancing on the spread of COVID-19. Starting in China, the virus has reached the United States of America with most numbers. Other countries severely affected by the pandemic are Brazil, Russia, United Kingdom, Spain, Italy, and France. Even though it is not possible to eliminate the spread of the virus from the world or any other country, however, it might be possible to reduce its effect by decreasing the peak of infected peoples. Implementing such policies needs a solid ground, which is impossible with a simple mathematical linear equation or Monte Carlo method because human society is a complex adaptive system with complex and continuous feedback loops. In such scenarios, an agent-based model is a good choice to investigate the action effects relationships.
2. Entities, State variables, and scales: This model contains only a single agent, which is a person. The person has three states: healthy, infected, and recovered. Agents are colored according to state and change their state according to the respective state. The healthy agents are colored with green, the infected are colored with red, and the recovered agents are colored with yellow. Each agent has one of two strategies: following social distancing or not following social distancing. Agents would move randomly on the canvas. As described above, the agents must have a label representing the strategy. “S” represents that agents are following “Social Distancing,” while the “N” represents that the agents are not following the social distancing policy. The environment is a 2D grid of 32×32 cells. One cell can contain only one agent at a time. Agents can randomly move inside the model, pursuing the strategy they consider feasible for survival.
3. Process Overview and Scheduling: Each agent must follow one of the two strategies: Social Distancing or Non-Social Distancing. A person moves randomly in the model, and encounter or meet another person. If the person is meeting another person who has COVID-19, then this healthy person would be affected by that sick person and get sick too. In the model, every person who is following the non-social distancing strategy would meet other persons. However, the persons who are following the social distancing strategy would not meet any others.

### B. Design Parameters

This section describes the design concept of the model.

1. Basic Principle: The basic principle of the model is to understand the effect of social distancing on COVID-19 spread. We would use two types of strategies to understand this effect.
2. Emergence: By employing the simple rule of social distancing, we found the emergent structure of healthy and sick people in society.
3. Adaption: People would adapt to the optimal strategy to remain healthy.
4. Objectives: The people’s objective is to remain healthy, and also maintain their relationships.
5. Learning: The agent would learn how to survive by following the social distancing rule.
6. Prediction: Overall death and survival rates.
7. Sensing: gents would sense the other agents who are infected with COVID-19.
8. Interaction: Every agent would interact with other agents randomly.
9. Stochasticity: Agent moves randomly in the canvas.
10. Collectives: There are no collectives in the model.
11. Observation: Infection, recovery, death, and survival rates are the observations.

### C. Details

1. Initialization: Every experiment has different values for initialization. The following figure depicts one of them. According to the Centers for Disease Control and Prevention records, the total infected and deaths cases are 6,706,374 and 198,099, respectively, until the moment [8]. So, the fatality rate is about 3%, and it would be used in all experiments as a fixed value. Also, Niklas Bobrovitz and his team at the University of Toronto assessed the disease test data to determine the proportion of infected people in the countries, where the infection rate is 6% in the United States [9].
2. Input Data: There is no input data in this model.
3. Sub Models:

**Table 1:** The experiment factors and levels

| No | Factor | Value |
| --- | --- | --- |
| 1 | Total People | 1000 |
| 2 | Infected people % | 6% |
| 3 | Fatality rate | 3% |
| 4 | Social distancing % | 0%, 30%, 40%, 60%, 80%, 100% |
| 5 | Canvas | 32X16 Grid |
| 6 | Social distancing label | Displaced as label |
| 7 | Color | Yellow = Recovered<br>Red = Infected<br>Green = Healthy |

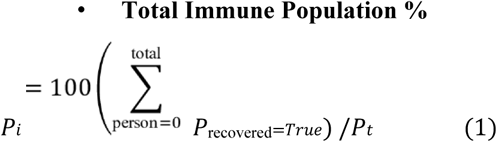

Where:

Pi is the total immune population Pt is the total population

Precovered=True is the total population recovered

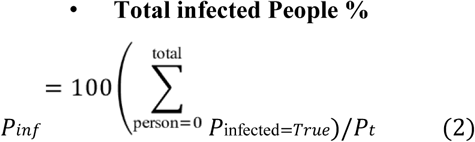

Where:

Pinf is the total infected population

Pt is the total population

Pinfected=True is the total population infected

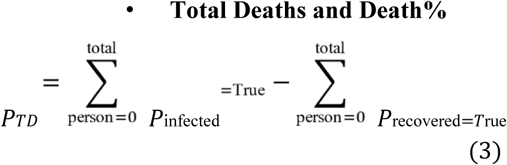

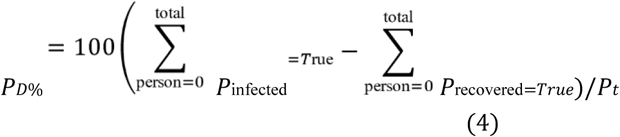

Where:

PD% is the percentage of total deaths from population

PTD is the total deaths

Pt is total the population

Pinfected=True is the total population infected

Precovered=True is the total population recovered

**Table 2:** The six experiments conditions/settings

| Experiment Number | Social Distancing Percent/ Level | Population | Infected Percent | Fatality rate |
| --- | --- | --- | --- | --- |
| 1 | 0% | 1000 | 6% | 3% |
| 2 | 30% | 1000 | 6% | 3% |
| 3 | 40% | 1000 | 6% | 3% |
| 4 | 60% | 1000 | 6% | 3% |
| 5 | 80% | 1000 | 6% | 3% |
| 6 | 100% | 1000 | 6% | 3% |

## III. RESULTS (GRAPHS, CHARTS, TABLES)

### A. Social Distancing 0%

In the first experiment, the social distancing percent slider was set up at zero percent to test the spread of COVID-19 without following social distancing procedures at all, as shown in Figure 6. Since we are just interested in studying and analyzing the effect of social distancing on the disease spread, the other factors values would remain fixed among all experiments such as the number of people in the sample, infected people percent, and fatality rate. In addition, the initial visualizations of all experiments tend to produce the same visual outputs, but the final visualizations would yield different visual outputs depending on the different percentages of followed social distancing in each experiment. Figure 7 shows the initial visualization of the first experiment where the infected and healthy people are represented by red and green agents, respectively. In addition, the disease transmission occurs through close contact between them; then, the infected cases would survive or die. So, the final visualization of the model gives a clear idea about the rates of healthy, infected, recovered, and deaths of people at the end of the experiment.

**Figure 1:**
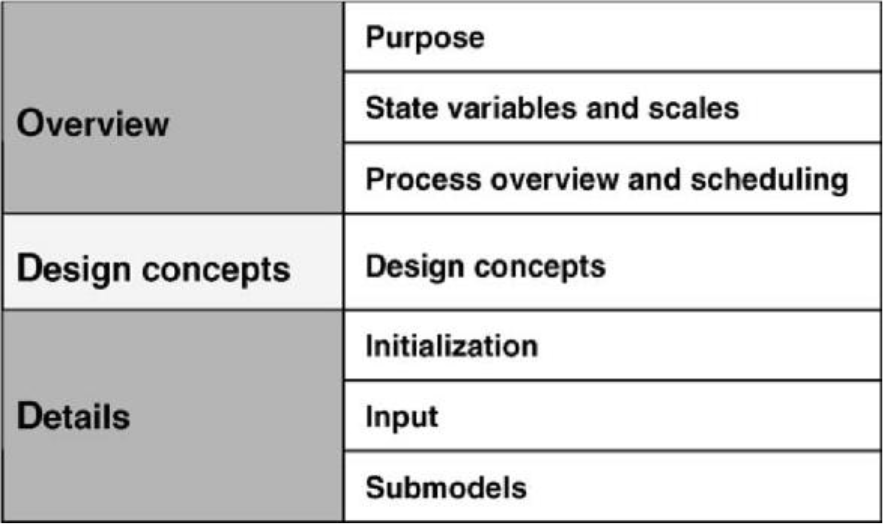
ODD Protocol.

**Figure 2:**
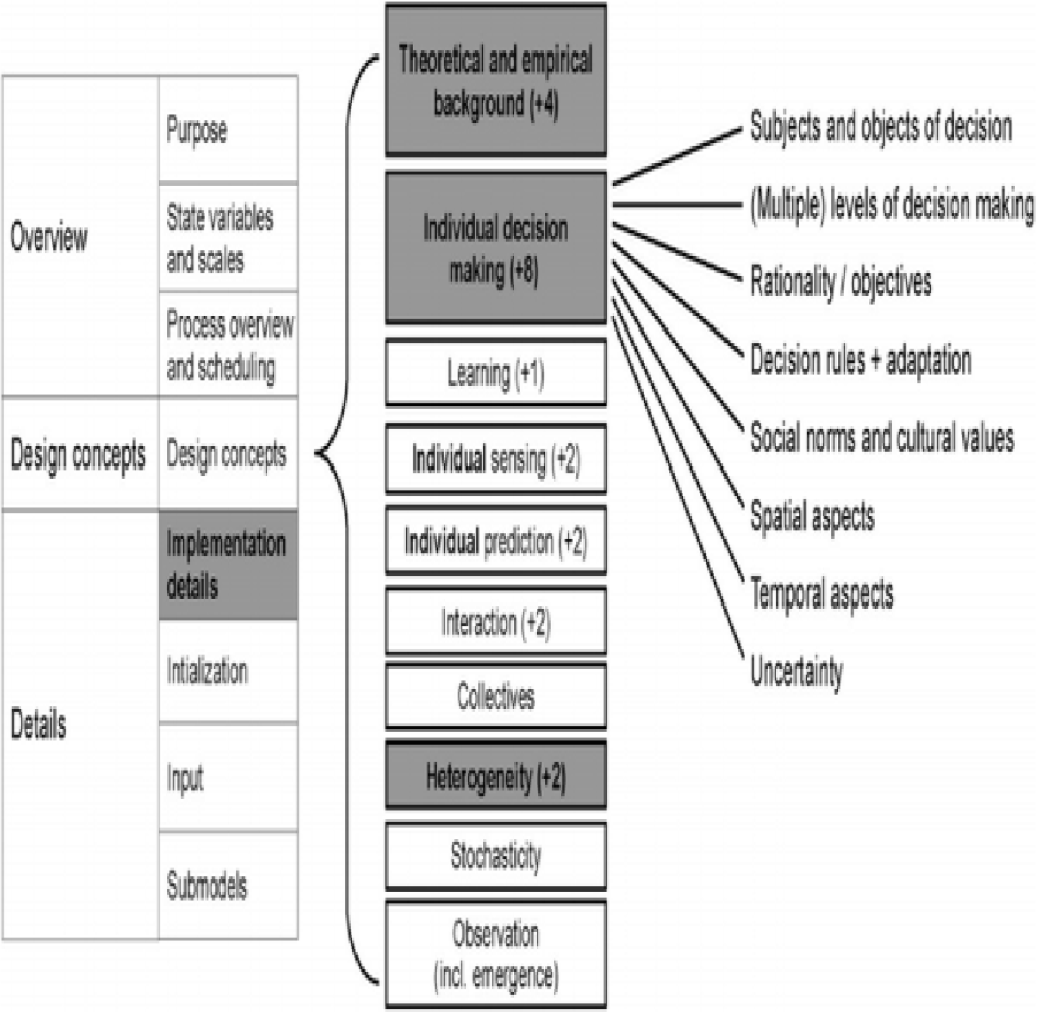
ODD+D protocol.

**Figure 3:**
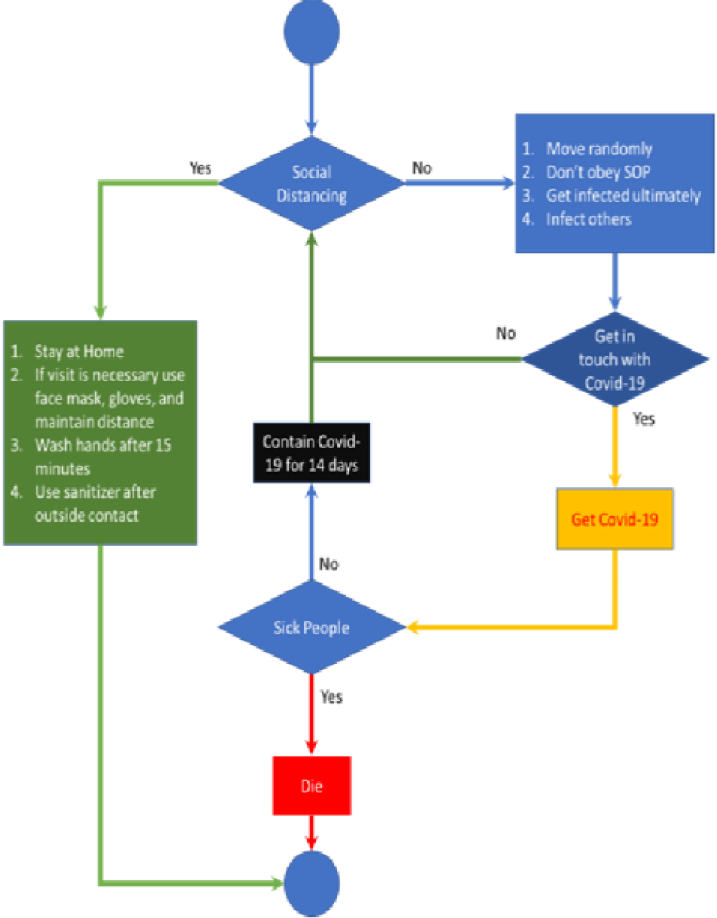
Process overview and scheduling.

**Figure 4:**
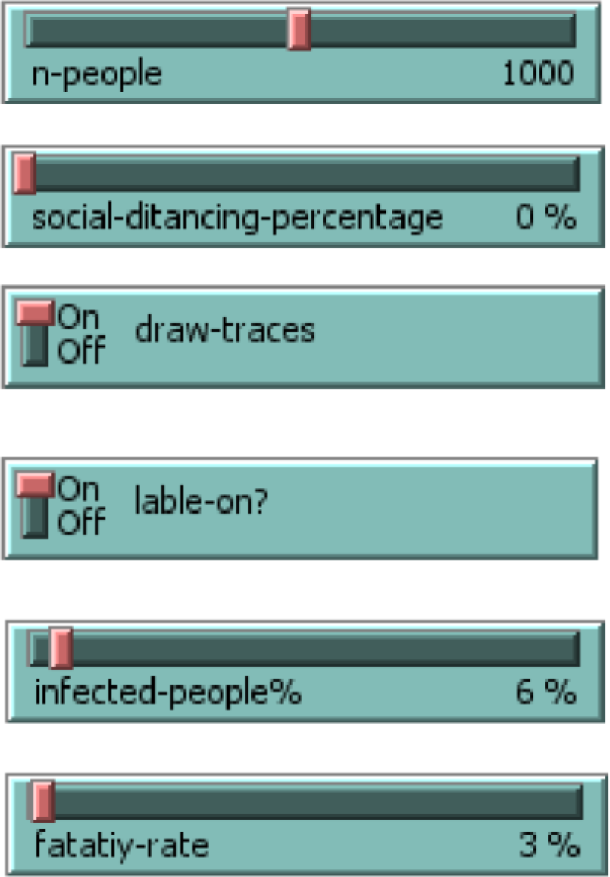
Sliders and switches.

**Figure 5:**
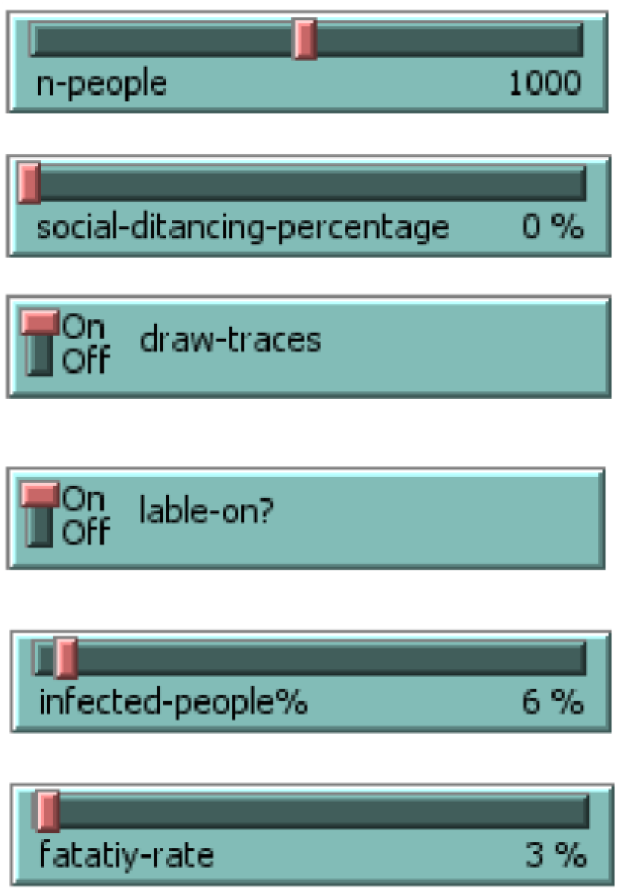
First experiment settings.

**Figure 6:**
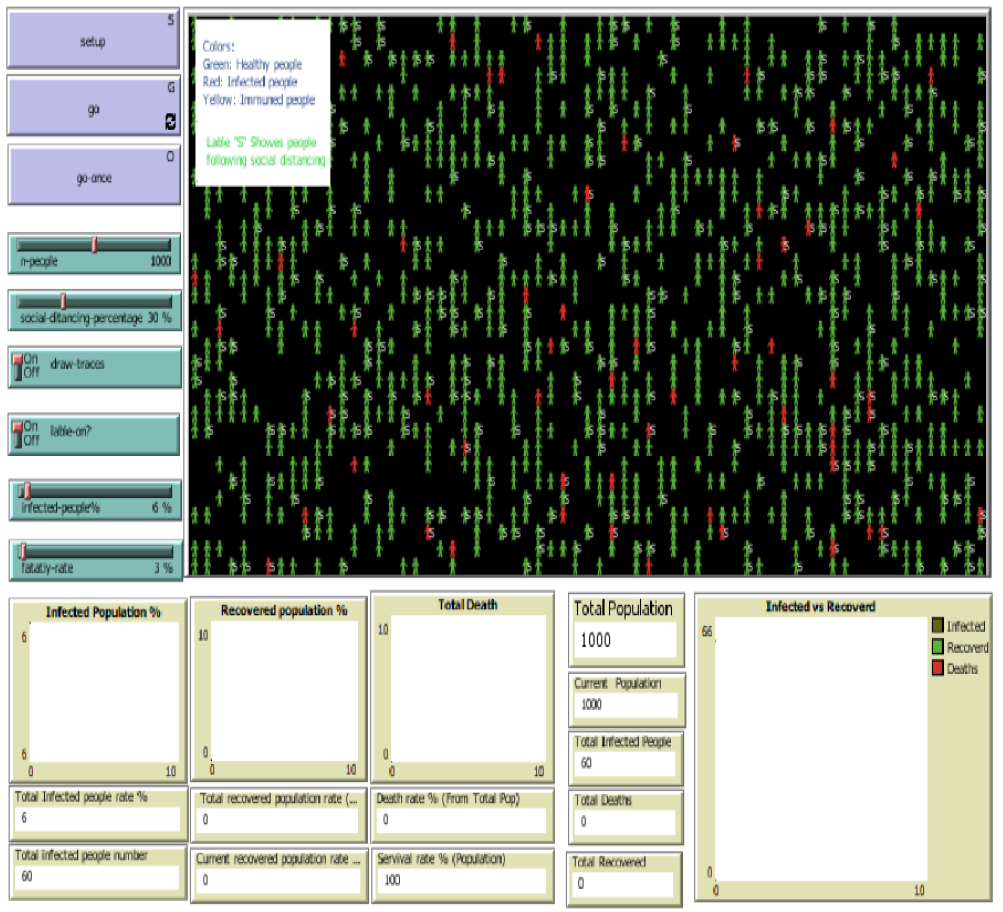
The Netlogo model interface.

**Figure 7:**
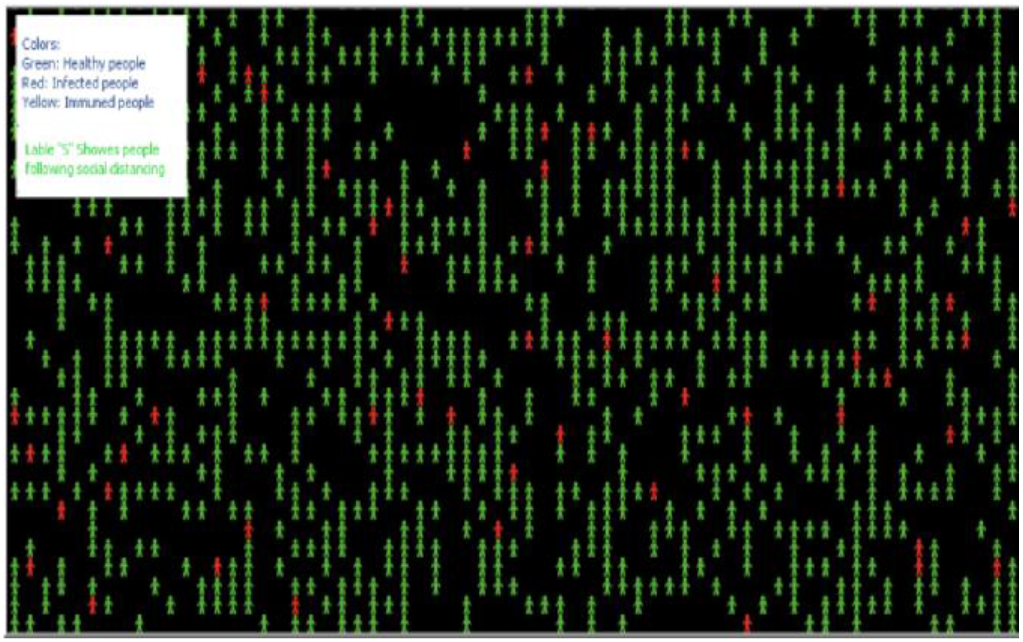
The initial visualization of the experiment.

After completing the first experiment, as shown in figures 8 and 9, it was feasible to read and analyze the final visualization and statistical records. To explain, the numbers of infected people were too high due to the absence of social distancing, and there were only 3% fatality rates. So, most of the infected cases have recovered, which explains why most of the agents have turned yellow at the end of the experiment, while the number of healthy people who have never infected is too small.

**Figure 8:**
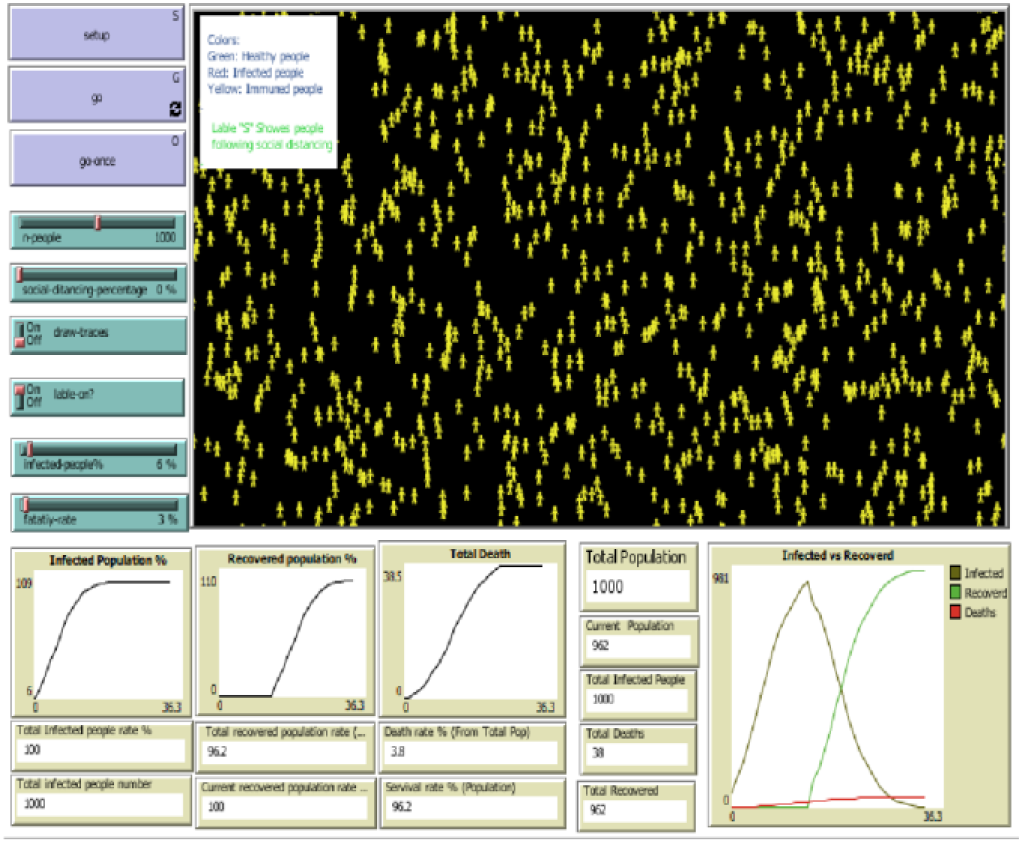
First experiment results.

**Figure 9:**
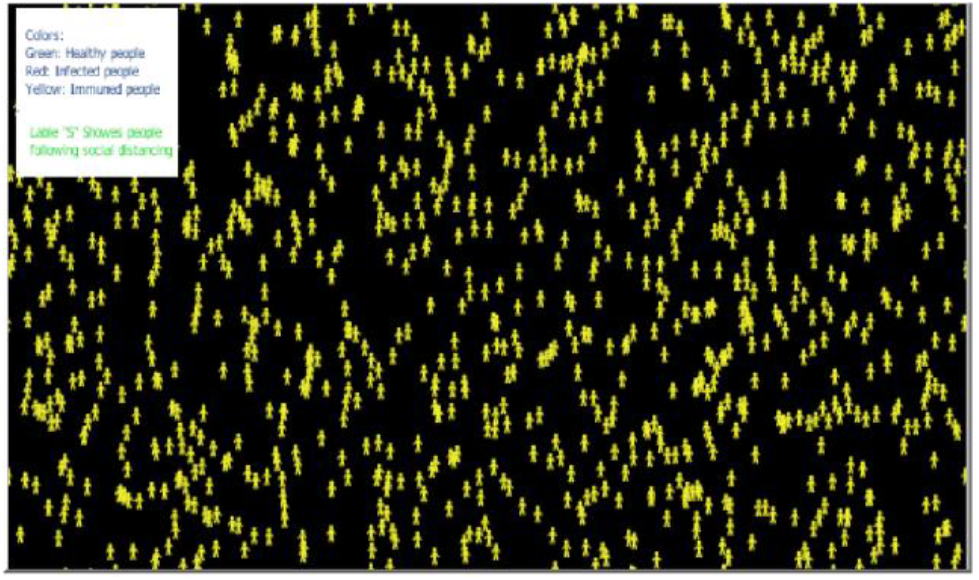
Final visualization of agent’s situation.

The movement patterns of the three different categories of agents were traced, as shown in figure 10, where the red, yellow, and green movement patterns represent the motion of infected, recovered, and healthy agents, respectively. It is clear that the infected and recovered people represent a huge part of the final visualization, while the health people represent a small part of motion patterns.

**Figure 10:**
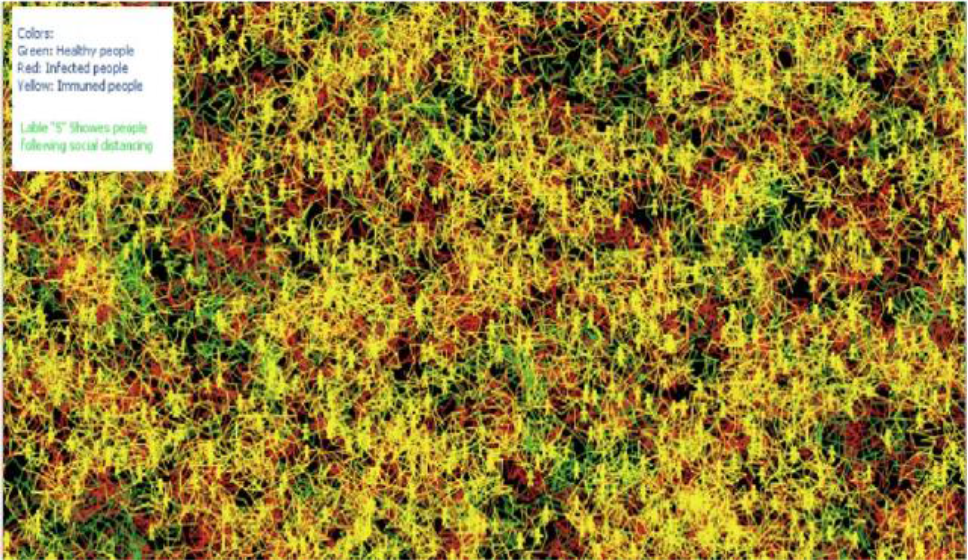
Movement patterns in the first experiment.

Figures 11 and 12 show the statistical estimates and records of total recovered, infected, and dead populations. These records are useful to compare all experiment results after using different levels of social distancing. It would also be possible to understand how changing social distancing strictness levels will influence the transmission of disease, which will allow decision-makers to create effective recommendations and regulations.

**Figure 11:**
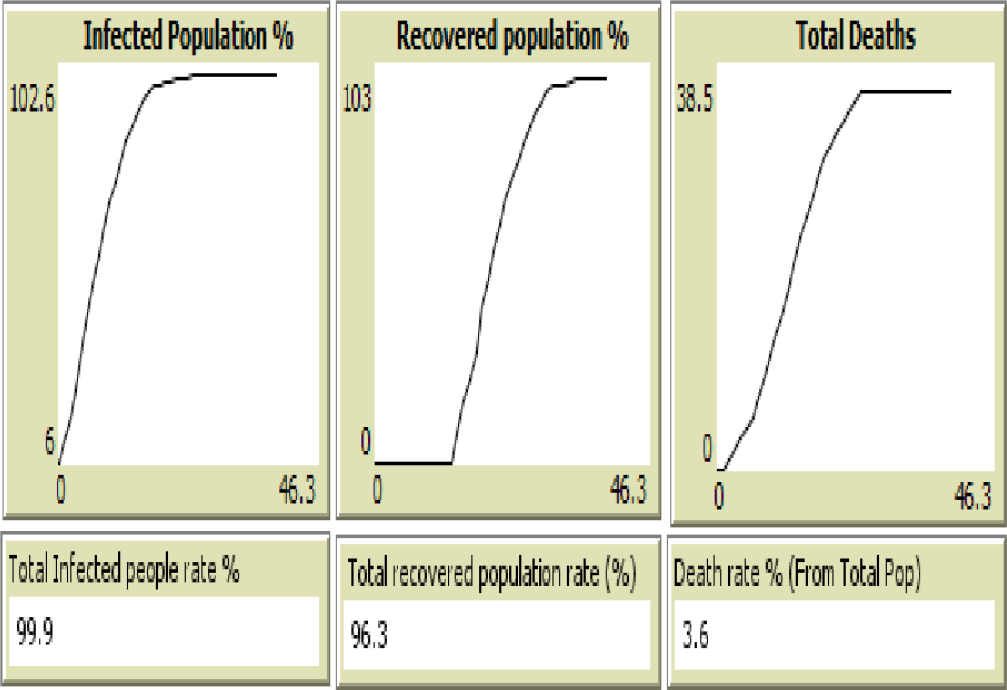
Infected, recovered, and deaths population rate.

**Figure 12:**
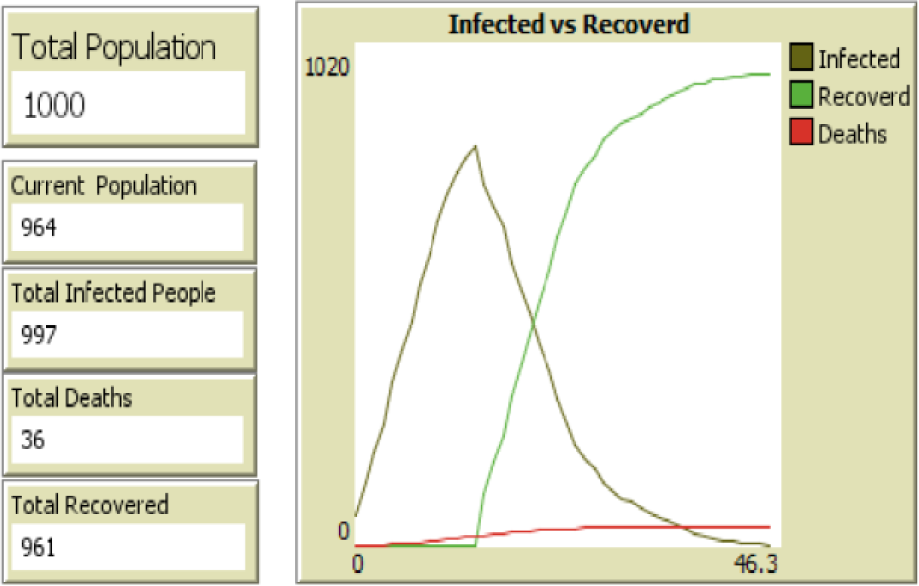
Infected versus the recovered people curve, when the fatality rate is 3%, and zero percent of social distancing.

### B. Social Distancing 30%

Then, the experiment was carried out with a higher level of social distancing, where the social distancing level was increased from zero to 30 percent without modifying any other parameters in the Netlogo model settings, as appears in figure 13.

**Figure 13:**
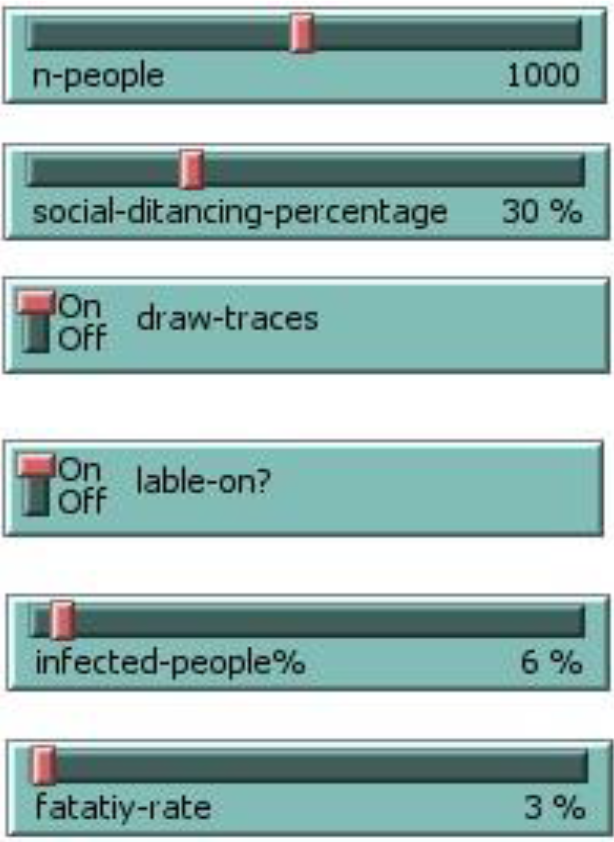
Second experiment settings.

The final visualization of movement patterns in the second experiment indicates that red and yellow patterns are lower compared to the first experiment, while the green patterns are greater in the second experiment, which means there is a significant improvement in reducing the probability of infection and increasing the survival rate.

Based on figure 15, the results of using 30\% of social distancing prove that the numbers of infected and deaths were reduced compared to the previous experiment. The number of infected people was 997 in the first experiment, but in the second experiment, it became 708 infected cases. Therefore, the decline in infected numbers of people in the second experiment lead to a decrease in the number of deaths too.

**Figure 14:**
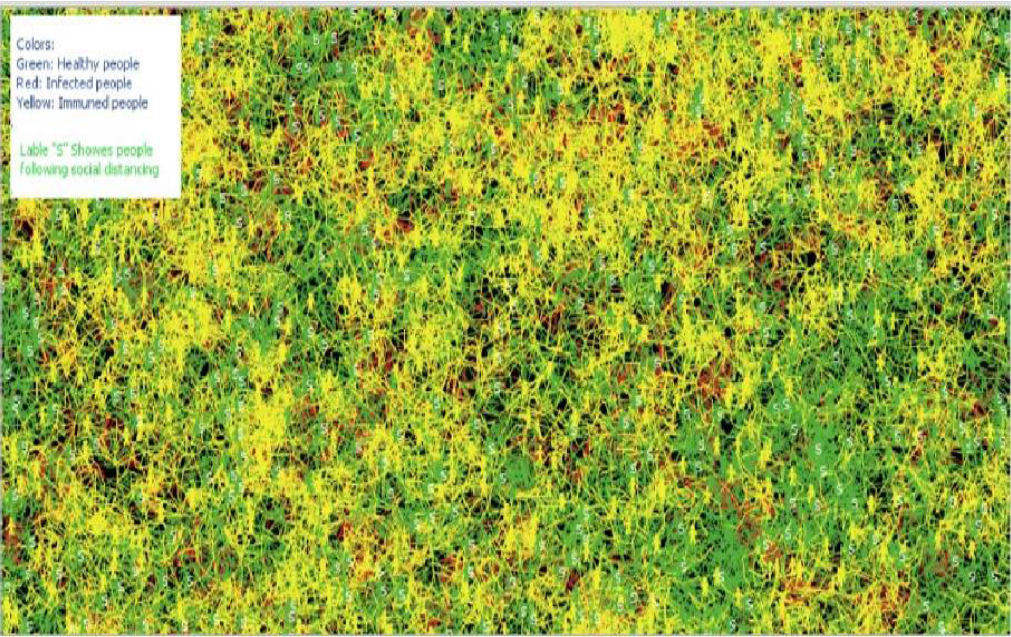
Movement patterns in the second experiment.

**Figure 15:**
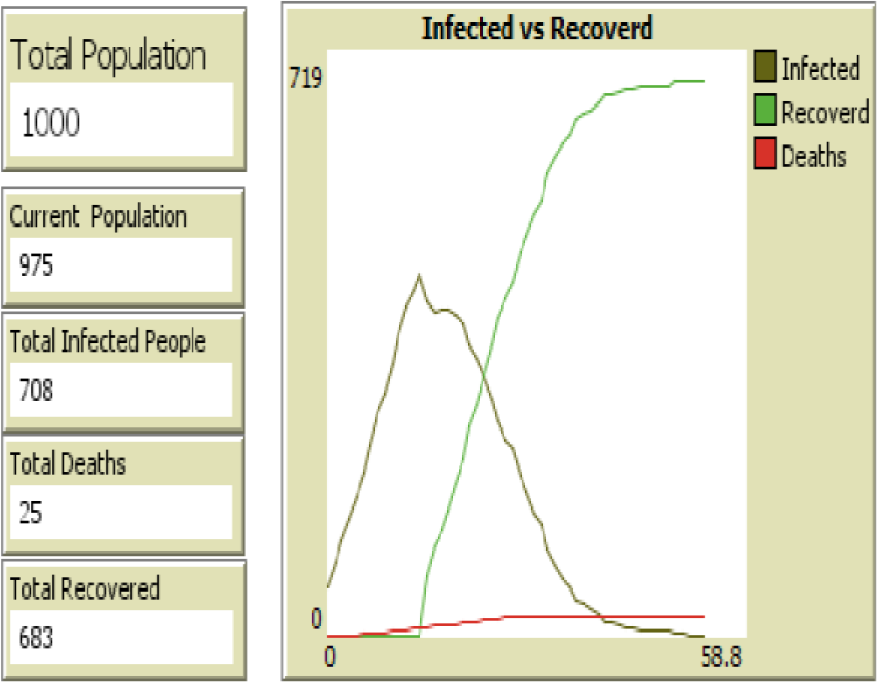
Infected versus the recovered people curve, when the fatality rate is 3%, and 30% percent of social distancing.

### C. Social Distancing 40%

Then, the percentage of social distancing was increased to the next level, which is higher and equal to 40%, resulting in further improvement and reduction in the spread of disease compared to the lower previous levels of social distancing, as shown in Figure 16.

**Figure 16:**
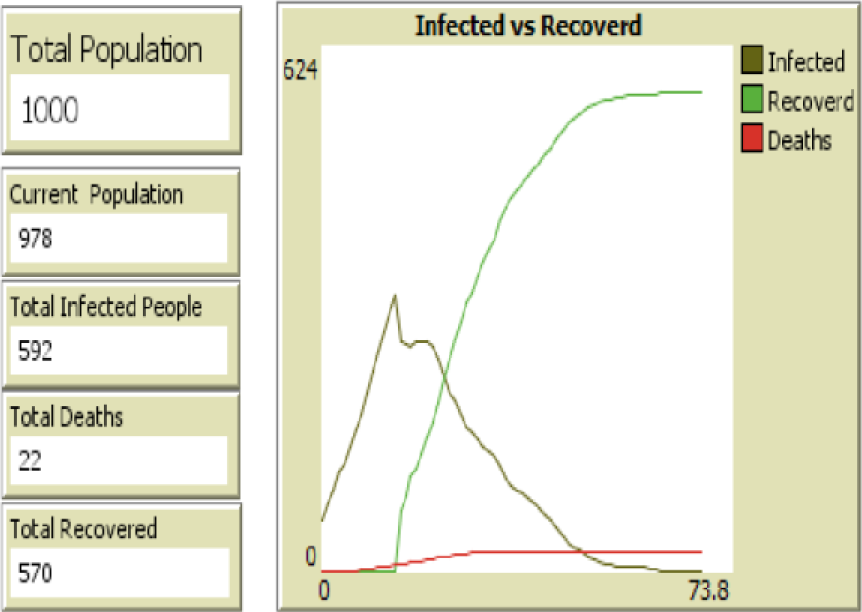
Infected versus the recovered people curve, when the fatality rate is 3%, and 40% percent of social distancing.

increasing the social distancing level to 60%, 80%, and 100% in the last three experiments, respectively. The final findings indicate significant improvements in protecting the population from infection, where the different visualizations of the six experiments are shown in figure 17.

**Figure 17:**
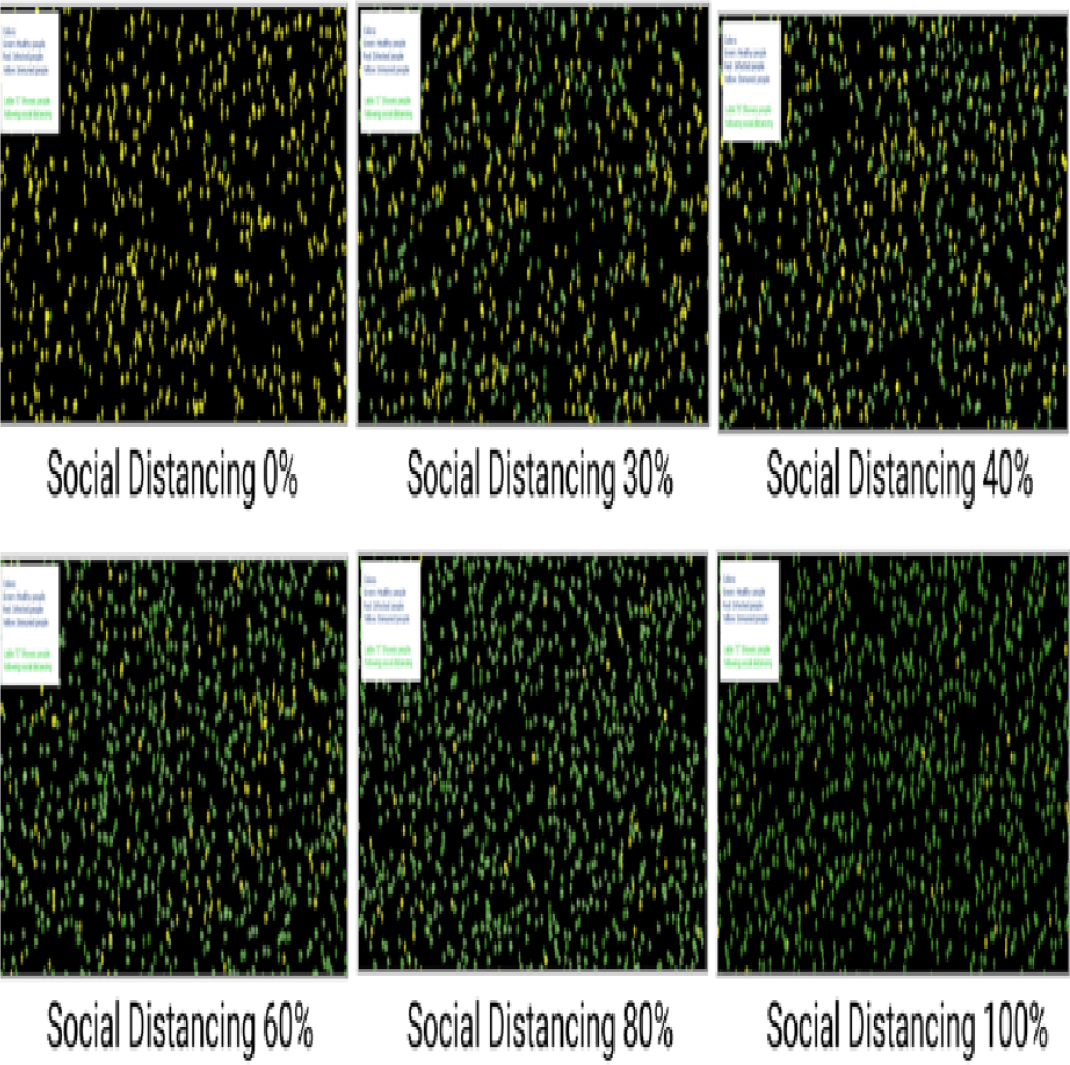
The visual outputs of applying different levels of social distancing.

By comparing the final visualizations of movement patterns of the six experiments, as shown in figure 18, we can see that increasing social distancing levels leads to reduce the spread of COVID-19 dramatically. To explain, following 100 percent of social distancing generated the best results with the highest rate of survival and the lowest rate of death.

**Figure 18:**
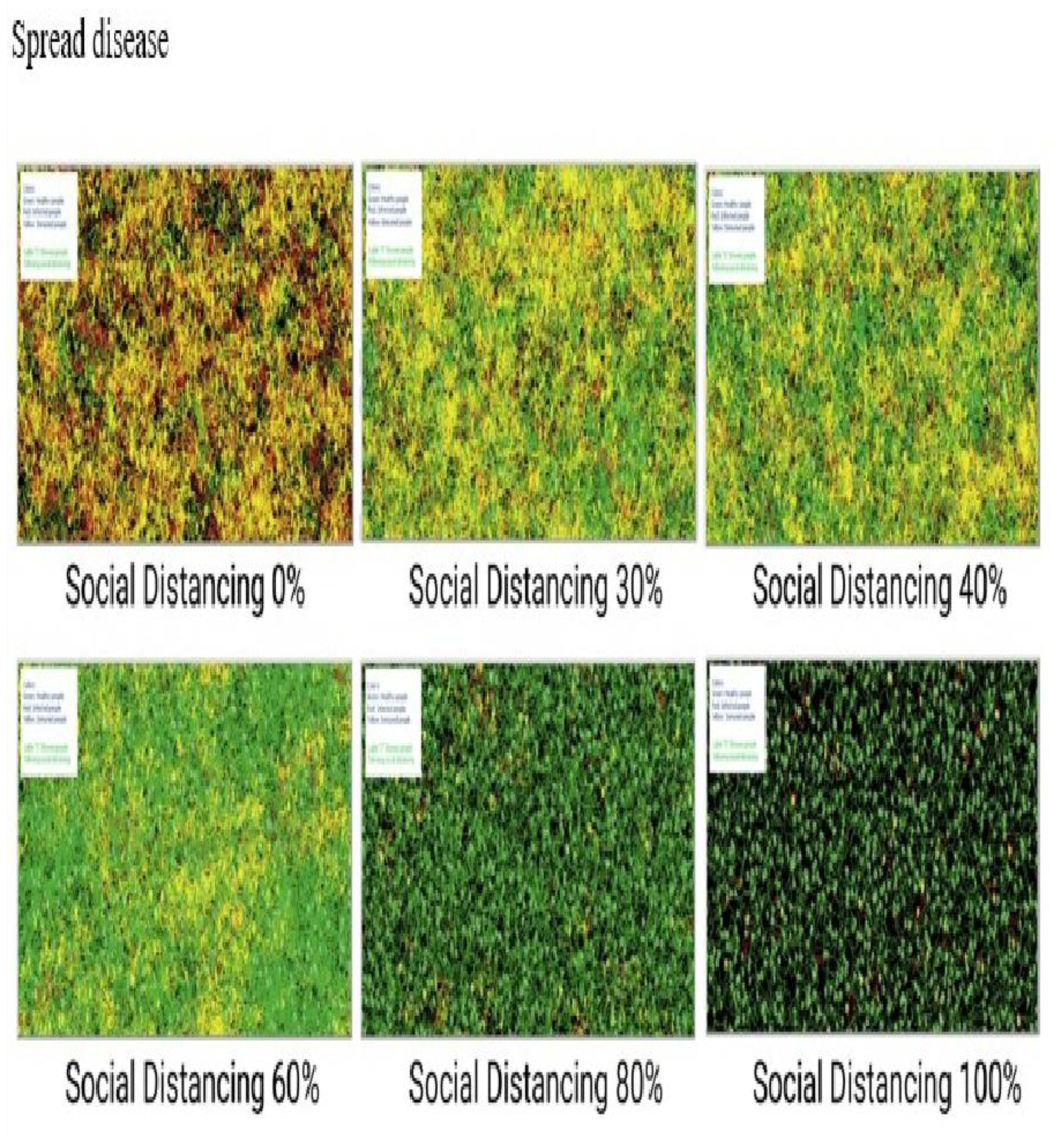
Final visualizations of movement patterns.

As we presented previously, the previous six experiments have visual outputs. However, there are ten other experiments done with ten different levels of social distancing by using the behavior space tool, and their results are shown in figure 19. The curve proved and confirmed that increasing the social distancing percentage leads to a lower infection rate, which results in decreasing the deaths and recovery rates.

**Figure 19:**
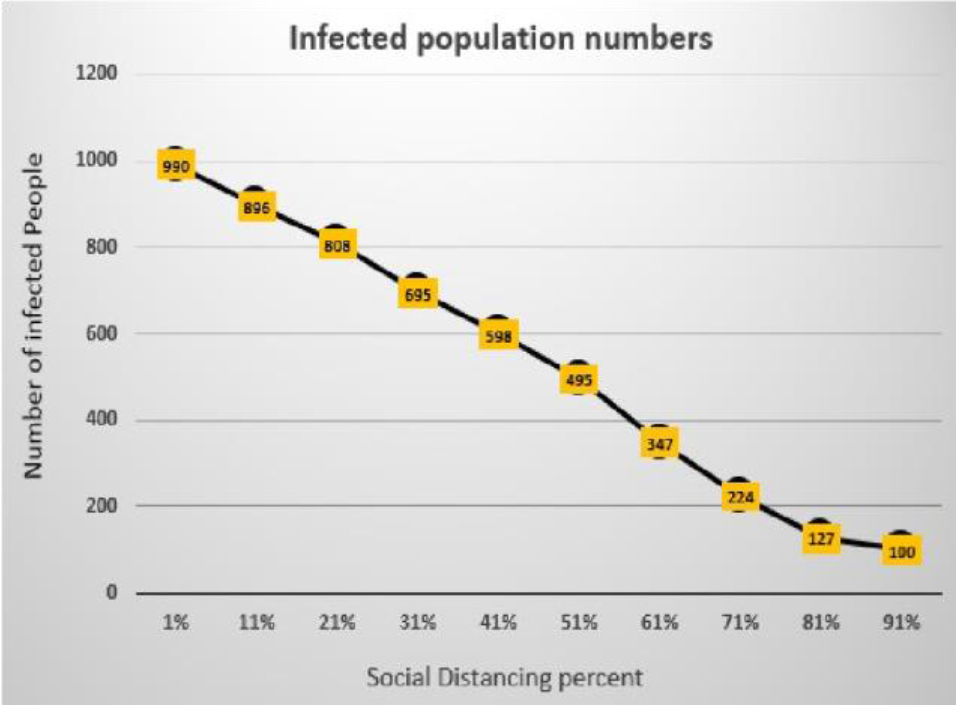
Effect of social distancing on infection rate.

The death rates and recovery rates are represented in figures 20 and 21, respectively, and they agree with what we discussed previously in figure 19. Since increasing the social distancing level reduced the infection rate initially, the number of deaths and recovery rates were declined too. In fact, the survival rate has been dramatically improved by analyzing the population’s survival rate, as shown in figure 22. Therefore, the study results support what the governments and agencies are doing right now about making strict instructions and procedures for keeping the social distancing phenomena exist everywhere in our daily lives until the disease disappeared completely.

**Figure 20:**
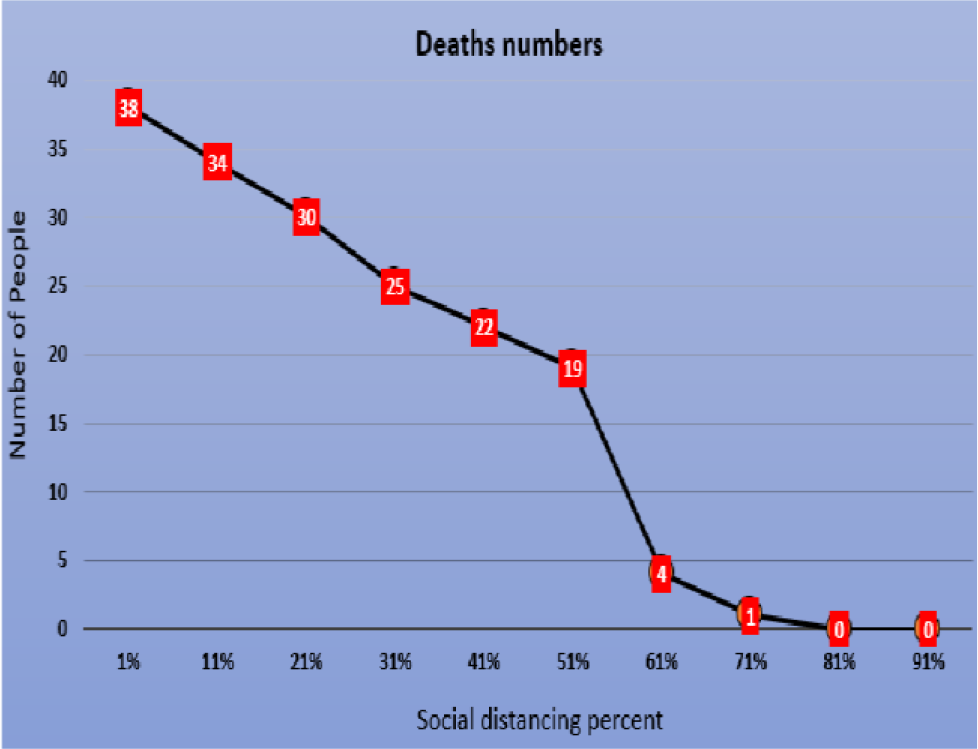
Deaths numbers.

**Figure 21:**
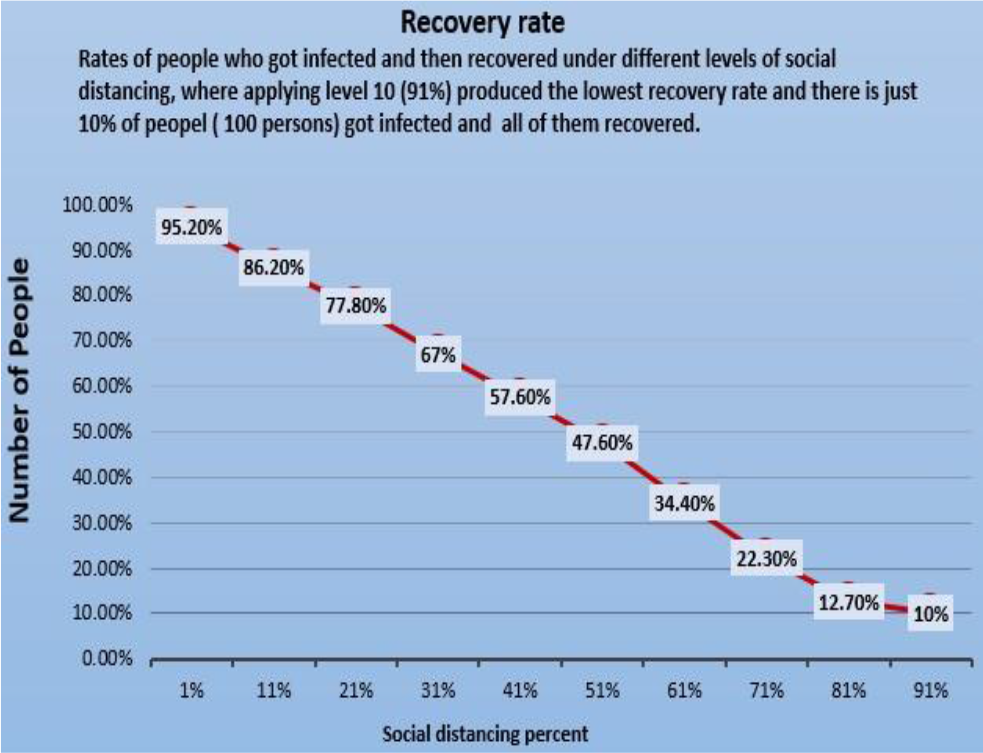
Recovery rate.

**Figure 22:**
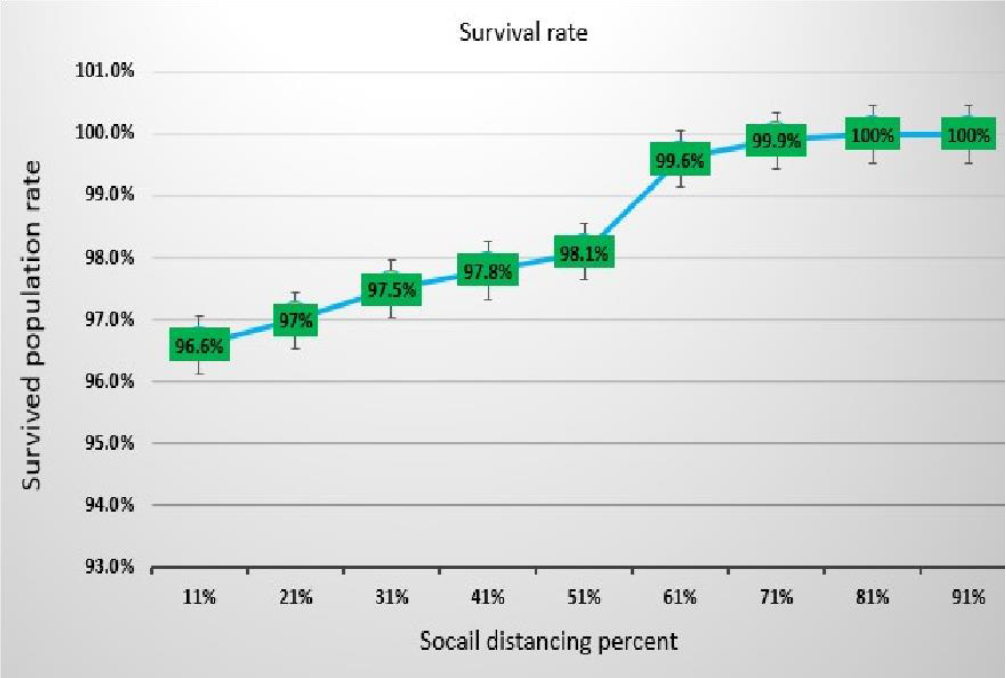
Survival rate.

## IV. DISCUSSION

In this agent-based simulation model, we verified that social distancing could help in reducing the spread of COVID19 and slow down the infection and fatality rates. We have observed with the help of simulation experiments that social distancing implication decreases infected cases, which helps in handling and treating severe patients. In other words, a lower number of critical cases means that the hospital team would be able to handle any case, so the chance of survival is far greater, and the healthcare system will never be stressed.

We have studied and simulated the social distancing behavior on discrete values ranging from zero to 100%. Therefore, we call it zero percent of social distancing when it is not implemented at all, and people do not follow any kind of preemptive measure to stop or reduce the spread of the pandemic. In the extreme case, we suppose that everyone is following the instruction to stop or reduce the spread of disease either implemented by law enforcement agencies or by the self-driving factors, and we call it 100% social distancing.

With the help of the experiment, it can be easily verified that the social distancing at zero percent implementation is very dangerous as compared to the social distancing at 80% to 100%. At zero percent of the social distancing, the spread of the COVID-19 is much faster, and the fatality rate is much higher as compared to the spread of disease at 100% social distancing. We can conclude that the enforcement of social distancing can help in reducing the spread of disease and also assist in handling the effected patient by the health care system very well.

One of the most prominent features that are really fascinating is that we can predict the outcome by using any materialistic resources and at a much lower cost. We can assist the decision-makers and legislators that what factors are more important to consider with the help of a replica of a real-world situation, which is not possible with the help of linear mathematical equations. Even though in agent-based simulation, very few variables are considered, the efficiency of the simulation model with respect to speed is not much satisfactory. However, with this small sampling, it can help in getting the overall picture of the system, which is very useful.

## V. Conclusions

In this model, we verified that social distancing affects the spread of COVID-19 and slows down its spread. It results in decreasing the active cases, which help in handling and treating serious patients. A few serious cases allow medical providers to handle each patient properly; thus, the chance of recovery is much greater. On the other hand, if the numbers of patients are higher, then we may not be able to accommodate and manage these cases in hospitals. In the United States, the level of social distancing intervention should be at least 80% to reduce the infected cases to the lowest number.

## VI. Future work

We are planning to extend the model to understand the effect of lockdown on the economy, which results in serious issues with the country’s overall growth rate, people’s survival, and unemployment. Our main purpose is to investigate how much lockdown can be eased so that the people would be able to earn enough money for survival and can be saved from the deadly virus.

## Data Availability

Yes

https://www.cdc.gov/coronavirus/2019-ncov/covid-data/forecasting-us.html

